# The 2020 U.S. cancer screening deficit and the timing of adults’ most recent screen: A population-based quasi-experiment

**DOI:** 10.1101/2022.06.24.22276836

**Authors:** Jason Semprini, Radhika Ranganthan

## Abstract

In 2020, cancer screenings declined, then rebounded, resulting in a cancer screening deficit. The significance of this deficit, however, has yet to be quantified from a population health perspective. Our study addresses this evidence gap by examining how the pandemic changed the timing of American adults’ most recent cancer screen. We obtained population-based, cancer screening data from the Behavioral Risk Factor Surveillance System. Mammograms, pap smears, and colonoscopies were each specified as a variable of mutually exclusive categories to indicate the timing since the most recent screening (never, 0-1 years, 1-2 years, 3+ years). Our quasi-experimental design restricts the sample to adults surveyed in January, February, or March. We then leverage a quirk in the BRFSS implementation and consider adults surveyed in the second year of each survey wave as the quasi-treatment cohort. Next, we constructed Linear and Logistic regression models which control for exogenous sociodemographic characteristics, state fixed effects, and temporal trends. Our results suggest that the deficit in 2020 was largely due to a one year delay among adults who receive annual screening, as the proportion of adults reporting a cancer screen in the past year declined by a nearly identical proportion of adults reporting their most recent cancer screen 1-2 years ago (3% to 4% points). However, the relative change was higher for mammograms and pap smears (17%) than colonoscopies (4%). We also found some evidence that the proportion of women reporting never having completed a mammogram declined in 2020, but the mechanisms for this finding should be further explored with the release of future data. Our estimates for the pandemic’s effect on cancer screening rates are smaller than prior studies, but because we account for temporal trends we believe prior studies overestimated the effect of the pandemic and underestimated the overall downward trend in cancer screenings across the country leading up to 2020.

## 1 Background

Despite the critical importance of early cancer detection, cancer screening rates in America are well below public policy targets, rife with racial and socioeconomic disparities, and despite considerable resources, stagnating (Suran 2022; Sabatino et al. 2022; Benavidez et al. 2021). Then came 2020, the first year of the COVID-19 Pandemic.

Even before the first wave of the COVID-19 Pandemic, predictions for cancer prevention and control systems were dire. Given the elevated risk to cancer patients, either from adverse COVID-19 outcomes or consequences of delayed cancer treatment, health systems needed to adapt to ensure the safety of cancer patients during these initial months of the pandemic (Al-Quteimat and Amer 2020; Anderson et al. 2020; Matos et al. 2021). These risk mitigation and continuity of care policies may have prevented dramatic declines in the proportion of cancer patients receiving care (Davidson et al. 2022; Semprini 2022). Unfortunately, cancer screening was less of a priority during the early months of the public health emergency (DeJong, Katz, and Covinsky 2021).

Cancer screening could have been impacted by a number of federal, state, or local public health emergency policies, as well as by the changing priorities and capacities of health systems, and by individual social distancing behaviors (Cancino et al. 2020; Richards et al. 2020; DeJong, Katz, and Covinsky 2021). More recent research illuminated the role of financial stress and time costs as individual-level barriers to cancer screening, both of which may have compounded the strain on health systems attempting to resume cancer screening programs (Hanna et al. 2022; Peoples et al. 2022; Findling, Blendon, and Benson 2020).

There has been no shortage of evidence highlighting the stark decline in cancer screening services during the initial stages of the pandemic. Evidence from hospital records or insurance claims have suggested that, compared to pre-pandemic levels, mammograms, pap smears and colonoscopies declined 60% to 90% in March/April 2020 (Duszak et al. 2020; Bakouny et al. 2021; Staib, Catlett, and DaCosta Byfield 2021. However, subsequent research has found that in the later months of 2020, claims and records of cancer screenings returned to pre-pandemic levels (Chen et al. 2021); DeGroff et al. 2021; Labaki et al. 2021; McBain et al. 2021; Bello, Chang, and Massarweh 2022; Drescher et al. 2022). Still, the pandemic created a major cancer screening deficit which may be difficult to address despite recent investment in “return to screening” initiatives (Chen et al. 2021; Hanna et al. 2022; Kelkar et al. 2022). Even as America returns to screening, some are being left behind (Mafi et al. 2022. In fact, among the adults reporting having delayed cancer screenings during the 2020 pandemic year only 25% have plans to return to screening (Suran 2022).

### 1.1 New Contributions

Clearly, the COVID-19 pandemic created a cancer screening deficit in 2020. However, our understanding of this deficit is limited to hospital and claims based data, which is not necessarily a valid representation of the population’s screening behavior. Moreover, the limited population-based research has attempted to quantify changing patterns in cancer screening by comparing rates in 2020 with 2019, or an average of prior years, essentially assuming that any change observed in 2020 was due to the pandemic (Fedewa et al. 2022; Dennis, Hsu, and Arrington 2021; Richardson 2022). This assumption, however, may not be valid as several factors could impact temporal screening patterns (Semprini 2022). Finally, few studies have attempted to infer the significance of this decline or deficit in cancer screenings in terms of how the time since the most recent cancer screen may have changed (Fedewa et al. 2022). This is critical for “return to screening” initiatives, as targeting or prioritizing health system capacity should consider the pandemic’s effect on the timing of a cancer screen; not just for adults who delayed care for a year, but for adults who delayed for longer or even delayed initiating their first cancer screen.

Our study addresses these evidence gaps by designing a population-based quasi-experimental Event History Analysis, where we compare the year-by-year change in the timing of the most recent cancer screen among adults not exposed to the 2020 pandemic with the change in adults exposed to a full year of the pandemic. This approach allows us to control for temporal trends in screening patterns and rigorously assess the internal validity of our design.

## 2 Methods and Materials

### 2.1 Data

We analyzed data from the Behavioral Risk Factor Surveillance System (BRFSS), a cross-sectional random-digit-dialed, telephonic survey (both landline and cellphone) of nationally representative sample involving non-institutionalized civilian population, aged 18 years or older, who reside in the United States (CDC 2021). This population based self-reported, ongoing survey is conducted across all 50 states, DC and three US territories, which collects information on behavioral health risks, chronic conditions and the usage of preventive services covering more than 400,000 adult interviews each wave year.

### 2.2 Sample

Our study population included non-institutionalized adults 18 years or older, residing in the US (including DC), interviewed between January 1st to March 31st, for the even years (2010 – 2020). Based on BRFSS cancer module eligibility, we included adults who were asked whether or not they have received a mammogram, pap smear and/or sigmoidoscopy/colonoscopy screening services (CDC 2021).

We then restricted our analysis to adults eligible for each respective cancer screen based on the United States Preventative Task Force: Mammograms (females age 40-74; Pap Smears (females age 25-64); and Colon/sigmoidoscopies (males and females age 45-79) (Moyer et al. 2012; Siu et al. 2016; Davidson et al. 2021). Participants from unknown US territories/jurisdictions, those who were interviewed between April 1st to December 31st for the years 2010 to 2020, those interviewed in the years which did not utilize cancer modules (odd years), were all excluded (Supplemental Figure 1).

### 2.3 Analytical Design

From the perspective of the analyst, the ideal experiment to evaluate how the COVID-19 pandemic impacted cancer screening would be to randomly assign adults into two groups: those exposed and those not exposed to the pandemic. After tracking individual screening behavior over time, any difference observed during the pandemic year (2020) would be attributed to the COVID-19 pandemic. This design is comically unrealistic. Unfortunately, quasi-experimental designs using “as-if” randomization into treatment and controls are also infeasible. Because the COVID-19 pandemic was a global event, everyone was exposed and everyone was impacted. Simple pre/post analyses have attempted to measure the change in cancer screening rates, but these approaches fail to account for other temporal factors which could be influencing screening rates in ways unrelated to the pandemic.

Rather than attempt to create different groups based on exposure to the pandemic during the pandemic year, we take a different approach by leveraging how BRFSS implements its cross-sectional survey over the course of 15 months. Selection into the BRFSS sample is random. When adults are surveyed, is also random. Therefore there is no reason to expect any observable differences in cancer screening behavior between adults queried early in a single year BRFSS survey wave and adults queried later in that same BRFSS survey wave; except however, during the COVID-19 pandemic year.

Our design begins by creating two distinct quasi-cohorts of adult BRFSS respondents. The first cohort only includes adults surveyed between January 1 and March 31 of the first year in each survey wave. We consider this “early” cohort the control group. Our second cohort only includes adults surveyed between January 1 and March 31 of the second year in each survey wave. We consider this “late” cohort the treatment group. Tables 2 (in the supplemental file) reports the cancer screening rates for both quasi-treatment and control cohorts for years 2010-2018. We test for significant differences in proportions between both groups and report the t-statistic and respective p-value to assess the comparability of these groups prior to 2020. We also conduct similar proportion tests for the sample composition of our control variables.

Prior to 2020, we hypothesize that screening behavior is not significantly different between our control and treatment groups. We also hypothesize that, prior to 2020, the trends in screening behavior do not differentially vary by group (see statistical analysis section below for details on these identification tests).

Evidence that levels and trends did not vary between groups prior to 2020 supports our identification assumption: that any difference in screening rates observed in 2020 should be attributed to the COVID-19 pandemic.

Statistical Analysis For each of the three cancer screenings (mammograms, pap smears, and colonoscopies), we model the probability of self-reported cancer screening behavior as mutually exclusive categories related to the timing of the most recent screen. This approach not only allows us to model the change in probabilities over time, but also model how the distribution of cancer screening behavior changes between each timing category. Our initial specification models the probability of a recent cancer screen as a series of linear probability models.

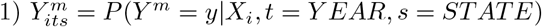

Where 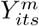 is a binary variable indicating if the respondent reported a cancer screening in time interval m. *X*_*i*_ is a vector of control variables potentially impacting cancer screening rates, but exogenous to the COVID-19 pandemic. This set of exogenous dummy variables includes age (five-year age groups), race/ethnicity (non-Hispanic White, non-Hispanic Black, Hispanic, and non-Hispanic other), education status (no highschool, high school degree only or GED, some college but no four-year degree, four-year college degree only or at least some graduate-level education and/or degree), whether the respondent is married, and whether the respondent is male. To account for secular trends in cancer screening, each model includes YEAR, a vector of binary fixed-effects variables indicating the survey wave year. These survey wave fixed effects account for temporal trends in cancer screening. Finally, to account for time-invariant, regional behaviors, policies, and health systems impacting cancer screening, each model includes s, a vector of binary fixed-effects variables indicating the state of residence for each respondent.

We then estimate the incremental effect of the COVID-19 pandemic on screening behavior 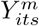 as:

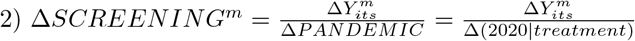

Where the estimated effect equals a change in 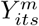 given an incremental change in pandemic status (modeled as the incremental change of a dummy variable indicating if the year is 2020, for adults in the treatment group). Rather than implement a simple pre/post design, we explicitly allow the screening behavior in the treatment group to vary from the screening behavior in the control group for each year of the analysis (Clarke and Tapia-Schythe 2021). The event history study model is defined as:

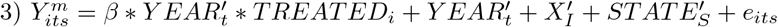

B identifies the average association between cancer screening behavior *Y* ^*m*^ for adults surveyed in the later part of each BRFSS survey wave. Using 2019 as the arbitrary baseline category, our event history analysis yields five B parameters (2010*Late, 2012*Late, 2014*Late, 2016*Late, and 2020*Late) This approach also allows us to test if 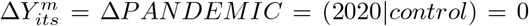 (Borusyak, Jaravel, and Spiess 2022). More importantly, we can now visually assess and empirically test our identification assumptions (Marcus and SantAnna 2021). Following best practice, we formally conduct pre-treatment differential trend tests by excluding responses in the 2020 survey wave and then recompute equation 3 (Freyaldenhoven, Hansen, and Shapiro 2019). In addition to reporting for each YEAR*TREATED coefficient, we calculate robust Wald statistics (with Bonferonni correction) to test if the trends in cancer screening rates between quasi-treatment and control cohorts jointly equalled zero (Armstrong 2014). Any significant results from these pre-treatment tests would suggest that cancer screening behaviors were differentially changing for treatment, compared to control cohorts, in ways unrelated to the COVID-19 pandemic. Conversely, null results provide confidence in the strength of our identifying assumption and supports the validity of our study design.

All analyses incorporate BRFSS supplied sampling weights, estimate standard errors robust to heteroskedasticity, and cluster the robust standard errors at the state level (Abadie et al. 2017; Cameron and Miller 2015). The Bonferonni correction method was used to adjust for multiple hypothesis testing for group differences and pre-treatment joint Wald tests (Armstrong 2014.

### 2.4 Alternative Specifications

Because of its ability to consistently model time-trends and group (i.e., state) fixed effects, the linear probability model is typically the preferred specification in the quasi-experimental literature. However, analytical issues may arise if the linear predictions are estimating potential outcomes outside the bounds of zero and one. In the presence of these nonsensical predictions, the linear probability model may be yielding biased estimates. To confront this threat, we construct an alternative set of specifications and model the probability of a recent cancer diagnosis with a multinomial logistic regression model. After comparing the predicted probabilities of these nonlinear models with the predictions of our linear model, we use the coefficients in the nonlinear models to estimate the average incremental effect of the treatment group (compared to the control group) in each year on the probability of each cancer outcome (Williams 2012). We also extend this marginal analysis to estimate the semi-elasticity, or relative change from baseline screening rates, for the treatment group in 2020 (Williams 2012). To support our inference, we test if the point estimates and standard errors in our linear models are significantly different from the estimates in the nonlinear specifications. A final set of sensitivity analyses relax the model assumptions by 1) removing the state-specific fixed effects, 2) remove state-specific fixed effects and estimate (unclustered) robust standard errors, 3) remove state-fixed effects, estimate (unclustered) robust standard errors, and ignore probability sampling weights.

## 3 Results

### 3.1 Event-History Estimates (Linear Model)

In short, we find evidence that in 2020, exposure to the COVID-19 pandemic was associated with changing patterns of self-reported cancer screenings. Our estimates reveal that for the quasi-treatment cohort (late survey wave) in 2020, reports of a cancer screen in the past year declined as reports of a most recent cancer in the past 1-2 years increased. In 2020 for the quasi-treatment cohort, we also estimate a significant decline in self-reports of never having had a mammogram, which corresponds to a 33% relative decline from baseline rates of women reporting never having completed a mammogram. The absolute change for reporting a most recent mammogram in the past 1-2 years increased 4.2% points, which corresponds to a 17.5% increase from pre-pandemic rates of having reported a recent mammogram in 1-2 years. Similarly, the absolute change for reporting a most recent pap smear in the past 1-2 years increased 4.2% points, representing a 17.3% relative increase from pre-pandemic rates. Finally, reports of a colonoscopy in the past year were estimated to have declined by 3.5% points, a 4.3% relative change from baseline.

### 3.2 Marginal Effect Estimates (Logistic Model)

Figures 1-3 visually report the non-linear, average marginal effect estimates (derived from the results of the multinomial logistic model). Here, for each screening type and category, the average marginal effect of belonging to the quasi-treatment cohort for each year of the analysis. For our 2020 year of interest, we estimate average marginal effects significantly different than zero for mammograms in the past 1-2 years, pap smears in the past 1-2 years, and colonoscopies in the past year. However, we also see years when the average marginal effect of belonging to the quasi-treatment cohort was significantly different than zero in pre-pandemic years.

**Figure 1:**
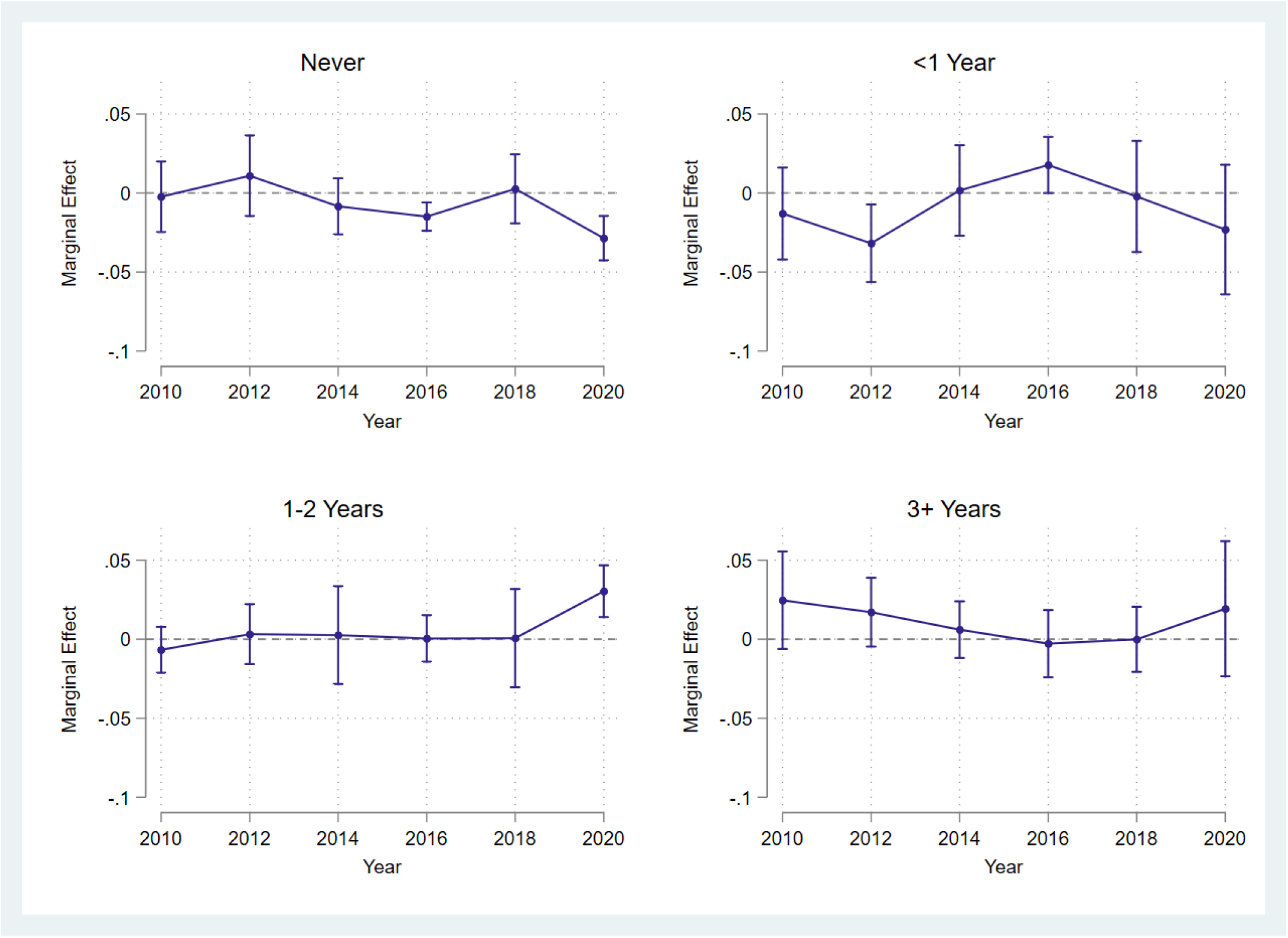
Marginal Effect of Late Survey Group on Rates of Most Recent Mammogram. Figure 1 shows the marginal effect estimates for each category of the most recent self-reported mammograms for the late-survey wave cohort. These estimates were based on the results of the multinomial logistic model and were averaged across all observations.

**Figure 2:**
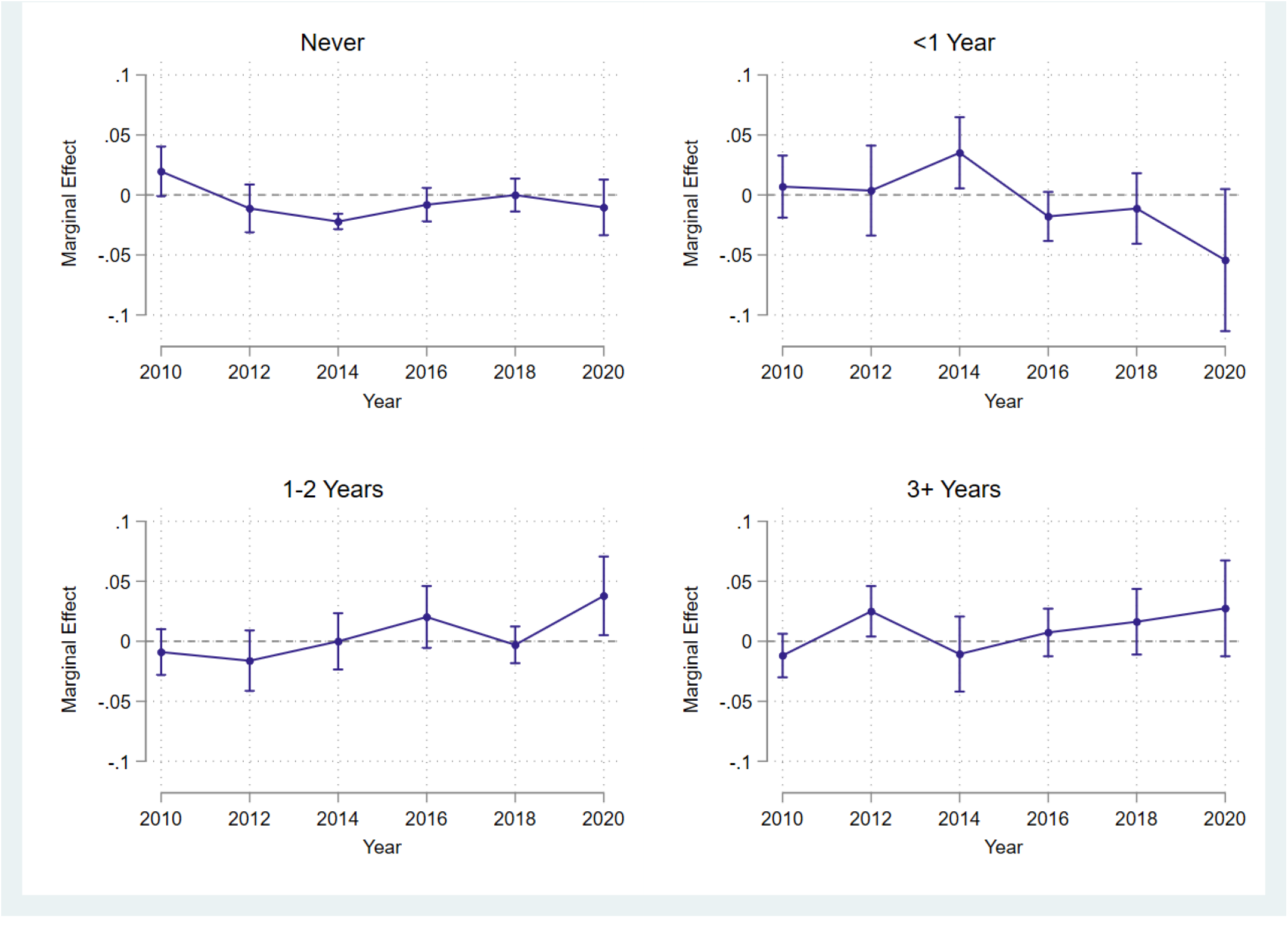
Marginal Effect of Late Survey Group on Rates of Most Recent Pap Smear. Figure 2 shows the marginal effect estimates for each category of the most recent self-reported pap smears for the late-survey wave cohort. These estimates were based on the results of the multinomial logistic model and were averaged across all observations.

**Figure 3:**
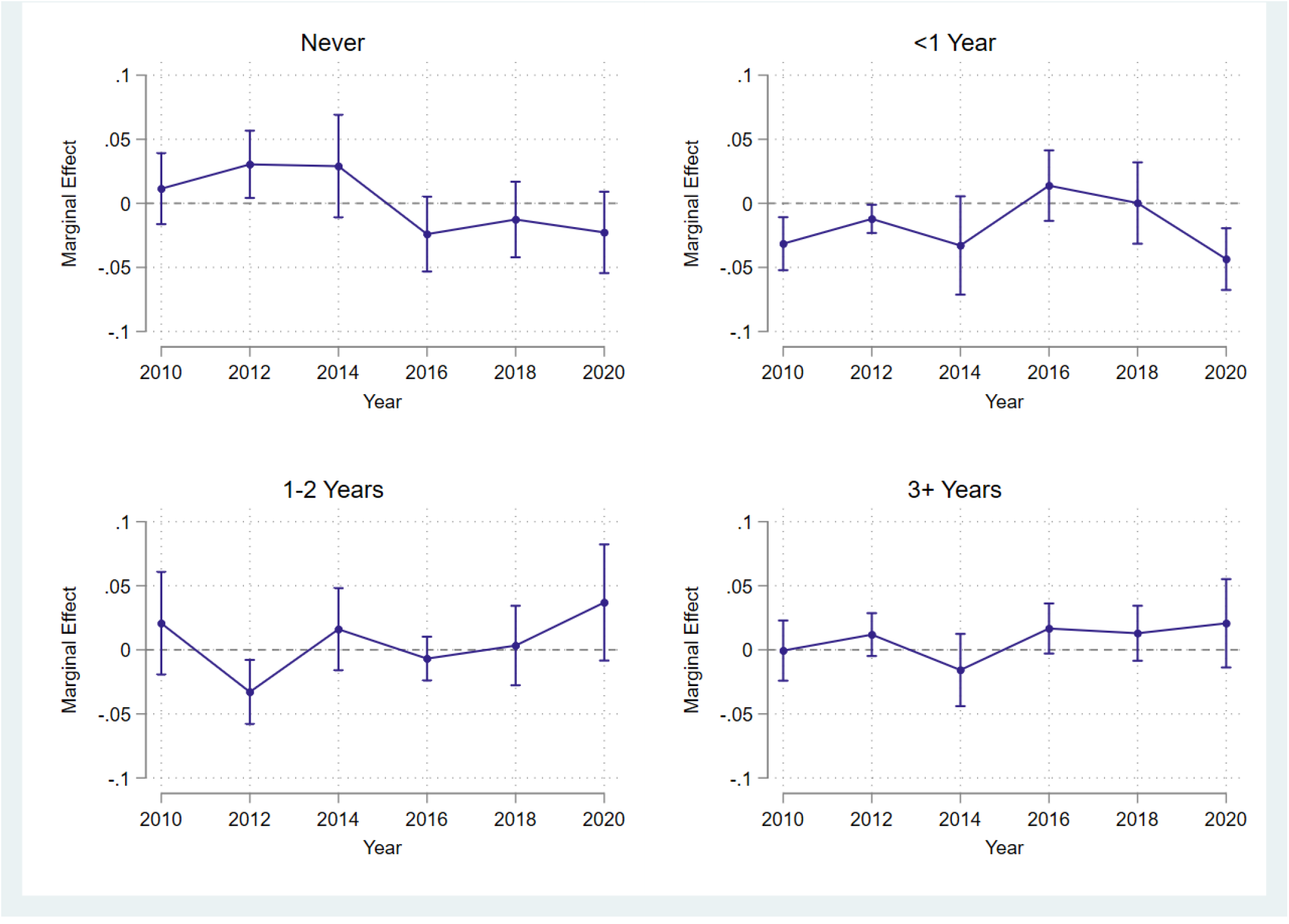
Marginal Effect of Late Survey Group on Rates of Most Recent Colon/Sigmoidoscopy. Figure 3 shows the marginal effect estimates for each category of the most recent self-reported colono-scopies for the late-survey wave cohort. These estimates were based on the results of the multinomial logistic model and were averaged across all observations.

### 3.3 Sensitivity Checks

Our effect estimates and inference do not appear sensitive to linear or non-linear model specification. Supplemental table 2 shows the 2020*Late effect estimates for the linear probability model specification and the multinomial logistic model specification. For each type of screening and screening category, none of the estimates are significantly different from each other. Further, our inference (whether to reject the null hypothesis that screening rates were similar in 2020 for the late survey cohort) changed. The major takeaway from Table 2 in the supplemental file is that the effect estimates in the non-linear model were more precise (smaller standard errors) for significant estimates for mammograms and colonoscopies.

Now while the estimates do not change with regard to including state-fixed effects, clustering robust standard errors, and weighting our analyses, our inference would change depending on the preferred specification. In the supplemental file, table 3 shows the alternative designs (Alt 1 - no state fixed effects, Alt 2 - no state fixed effects, no clustering robust standard errors, Alt 3 - no state fixed effects, no clustering robust standard errors, and no probability sampling weights). For each of the three alternative specifications, the effect estimates for having never reported a mammogram in 2020 lose their significance. Moreover, for effect estimates of mammograms in the past year and past 1-2 years, inference appears to change across alternative specifications. Inference for the effect estimates on pap smears and colonoscopies also, slightly, varies across alternative specifications. However, more interest were the increased number of significant pre-trend test statistics when excluding state fixed effects for both types of screenings. We suggest that this finding warrants inclusion the of the state-fixed effects to account for differential, time-invariant screening rates between quasi-treatment cohorts. Moreover, failing to account for clustering and weighting within each state may also add noise to our inference. Compared to our primary specification, our sensitivity analyses result in more differential pre-trend tests where we reject the null hypothesis (of no differential trends). Thus, we conclude that our primary specification is the most internally valid design.

### 3.4 Assessing internal validity

#### 3.4.1 Comparability of quasi-treatment cohorts

Upon observing the unadjusted reports of a most recent cancer screen prior to 2020, we find little convincing evidence to believe that screening rates in the two groups differed before the first pandemic year. Supplemental Figures 2-4 visually depict the unadjusted cancer screening rates for our quasi-treatment (late survey wave) and quasi-control (early survey wave), for each year in our analyses. For each category of mammograms and pap smears, we see nearly identical trends and levels in the most recent screening from 2010-2018. For adults reporting never having completed a colonoscopy and adults reporting a colonscopy in the past year, we do see some possible non-common levels from 2010-14, but the trends and levels appear similar from 2016-2018.

The results of our two-sample proportion tests further validate our assumption, that these two quasitreatment cohorts had simliar baseline screening rates. Note that Table 4 in the supplemental files does report a few screening outcomes with p-values under 0.05, but these test statistics are not significantly different than zero after accounting for multiple hypothesis testing (significance threshold p ¡ 0.0125). Further, we fail to reject the null hypothesis that the sample composition of these two quasi-treatment groups are significantly different from each other (Table 5 in supplemental files).

#### 3.4.2 Differential Trend Tests

We found evidence that screening rates in 2020 were significantly different between the two quasi-treatment cohorts. To infer a causal effect, however, we rely on the assumption that, in the absence of the pandemic, the change in screening rates would not have been observed for the late-survey wave cohort. To test the validity of this assumption, we empirically tested for differential trends prior to 2020. In the supplemental file, table 6 reports the year-by-year effect estimates (absolute differences) between quasi-treatment cohorts after excluding year 2020 from the analysis. We find no statistically significant differences in these effect estimates for mammograms. Moreover, the joint test of significance for each mammogram category exceeds the threshold to reject the null hypothesis that 2010, 2012, 2014, and 2016 coefficients equal zero (note, multiple hypothesis testing threshold set at p ¡ 0.0125).

We do however, detect the potential for differential trends in pap smears, specifically for adults reporting never having completed a pap smear and having completed a pap smear in the past year. The source of the potential pre-trend differences are observed in 2014. For both categories, we reject the null hypothesis that pap smear screening trends were similar prior to 2020 (p ¡ 0.0125).

Additionally, we detect the presence of differential trends for adults reporting their most recent colon/sigmoidoscopy three or more years prior. Again, the source of the differential pre-trend is observed in 2014. We reject the null hypothesis that, prior to 2020, reports of a colon/sigmoidoscopy three or more years ago were similar between the two quasi-treatment groups (p ¡ 0.0125).

## 4 Discussion/Conclusion

The dire predictions and early evidence that cancer screening dramatically declined prompted investment and capacity for “return to screening” initiatives and patient prioritization policies (Joung et al. 2022; Sprague et al. 2021). However, most early predictions focused on the initial decline, as opposed to the subsequent rebound, and so did most of the early research. Even the research which accounted for the rebound, failed to account for other, non-pandemic factors which could be biasing the cancer screening deficit estimate. Our study does not claim that the pandemic had no impact on cancer screening rates over the course of 2020. We found higher proportions of women completing their most recent mammogram and pap smear 1-2 years ago. We also found lower proportion of adults completing their colonoscopy in the past year. However, our point estimates are smaller than the most recent population-based research (Fedewa et al. 2022). We attribute this difference to the fact that our study includes adults exposed, not just to the entire pandemic year, but to the “rebound period”. Additionally, our Event-History design attempts to control for temporal changes which could be affecting cancer screening rates in ways unrelated to the pandemic, which other studies may fail to identify with simple pre/post designs. The implications of our findings suggest that “return to screening” initiatives and prioritization policies based on the overestimated effects of the pandemic on screening, may fail to achieve greater screening adherence. This is especially true for adults who have been delaying recommended cancer screenings for three or more years, delays which started before the pandemic year. To advance cancer equity, future research must continue monitoring the post-2020 cancer screening rebound to assess who is still delaying cancer screenings and how effective programs are mitigating these long-term delays. Finally, our results not only signal delayed cancer screening in 2020, but increased initiation. The proportion of women reporting to have never completed a mammogram declined for our late cohort in 2020 (relative to the change in the early cohort). Did the “return to screening” policies navigate new patients to their first mammogram? Or, was this decline merely a result of a “lower population denominator” after a year of elevated excess mortality? Future data can help us understand the mechanisms influencing the post-COVID screening rebound, which will be critical for advancing efforts to improve early cancer detection in America well beyond this pandemic.

**Table 1:** Year-by-Year Differences in Screening Rates for Late Survey Cohort

|  |  | 2010*Late |  | 2012*Late |  | 2014*Late |  | 2016*Late |  | 2020*Late |  | 2020 Relative Change |
| --- | --- | --- | --- | --- | --- | --- | --- | --- | --- | --- | --- | --- |
| Mammogram | Never | -0.006 | (0.018) | 0.009 | (0.016) | -0.013 | (0.014) | -0.017 | (0.012) | -0.029* | (0.013) | -33.0% |
|  | <1 Year | -0.011 | (0.023) | -0.030 | (0.024) | 0.004 | (0.023) | 0.020 | (0.021) | -0.021 | (0.034) | -3.5% |
|  | 1-2 Years | -0.008 | (0.020) | 0.003 | (0.018) | 0.002 | (0.020) | -0.000 | (0.016) | 0.031* | (0.015) | 17.5% |
|  | 3+ Years | 0.025 | (0.021) | 0.018 | (0.018) | 0.007 | (0.012) | -0.002 | (0.015) | 0.020 | (0.024) | 11.5% |
| Pap Smear | Never | 0.017 | (0.010) | -0.011 | (0.014) | -0.024* | (0.009) | -0.009 | (0.010) | -0.011 | (0.012) | -22.2% |
|  | <1 Year | 0.019 | (0.023) | 0.015 | (0.025) | 0.046* | (0.020) | -0.007 | (0.015) | -0.043 | (0.031) | -13.5% |
|  | 1-2 Years | -0.008 | (0.015) | -0.014 | (0.017) | 0.003 | (0.017) | 0.024 | (0.014) | 0.042* | (0.019) | 17.3% |
|  | 3+ Years | -0.028 | (0.015) | 0.010 | (0.019) | -0.026 | (0.017) | -0.008 | (0.017) | 0.012 | (0.018) | 10.2% |
| Colonoscopy | Never | 0.025 | (0.017) | 0.045* | (0.021) | 0.042 | (0.029) | -0.011 | (0.022) | -0.010 | (0.020) | -1.06 |
|  | <1 Year | -0.032 | (0.022) | -0.012 | (0.017) | -0.031 | (0.030) | 0.013 | (0.018) | -0.035* | (0.016) | -4.3% |
|  | 1-2 Years | 0.016 | (0.031) | -0.035 | (0.020) | 0.011 | (0.025) | -0.009 | (0.019) | 0.030 | (0.016) | 3.8% |
|  | 3+ Years | -0.009 | (0.020) | 0.002 | (0.012) | -0.023 | (0.019) | 0.008 | (0.018) | 0.015 | (0.019) | 2.1% |

Table 1 reports the year-by-year differences in cancer screening rates between the early and late survey cohorts. All analyses control for exogenous sociodemographic characteristics, state-level fixed effects, and temporal trends (year fixed effects). Robust standard errors were clustered at the state level and reported in parentheses. The relative change was estimated by computing the semi-elasticity (relative marginal effect) of belonging to the late survey cohort in 2020. \* $p < 0.05$ .

## Data Availability

BRFSS data is publicly available from CDC. The analytical code to create the dataset, clean/code variables, and replicate the analysis are included in this submission.

https://github.com/jsemprini/cancerscreening_covid_BRFSS/blob/main/BRFSS_CancerScreen_ReplicationFile.do

