## Supplemental Tables 1-6 for "The 2020 U.S. cancer screening deficit and the timing of adults’ most recent screen: A population-based quasi-experiment"

### 7 Supplemental Files

#### 7.1 Supplemental Figure 1: Sample Selection Flow Chart

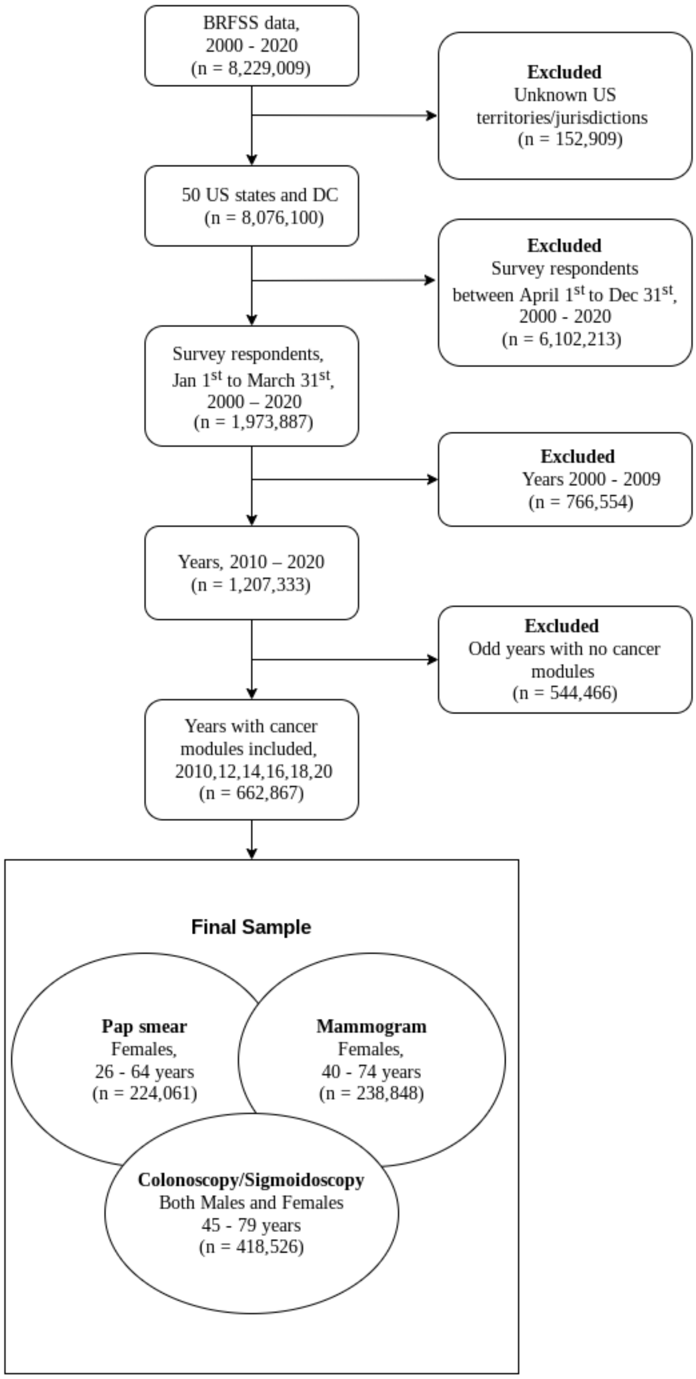

#### 7.2 Supplemental Figure 2: Mammograms

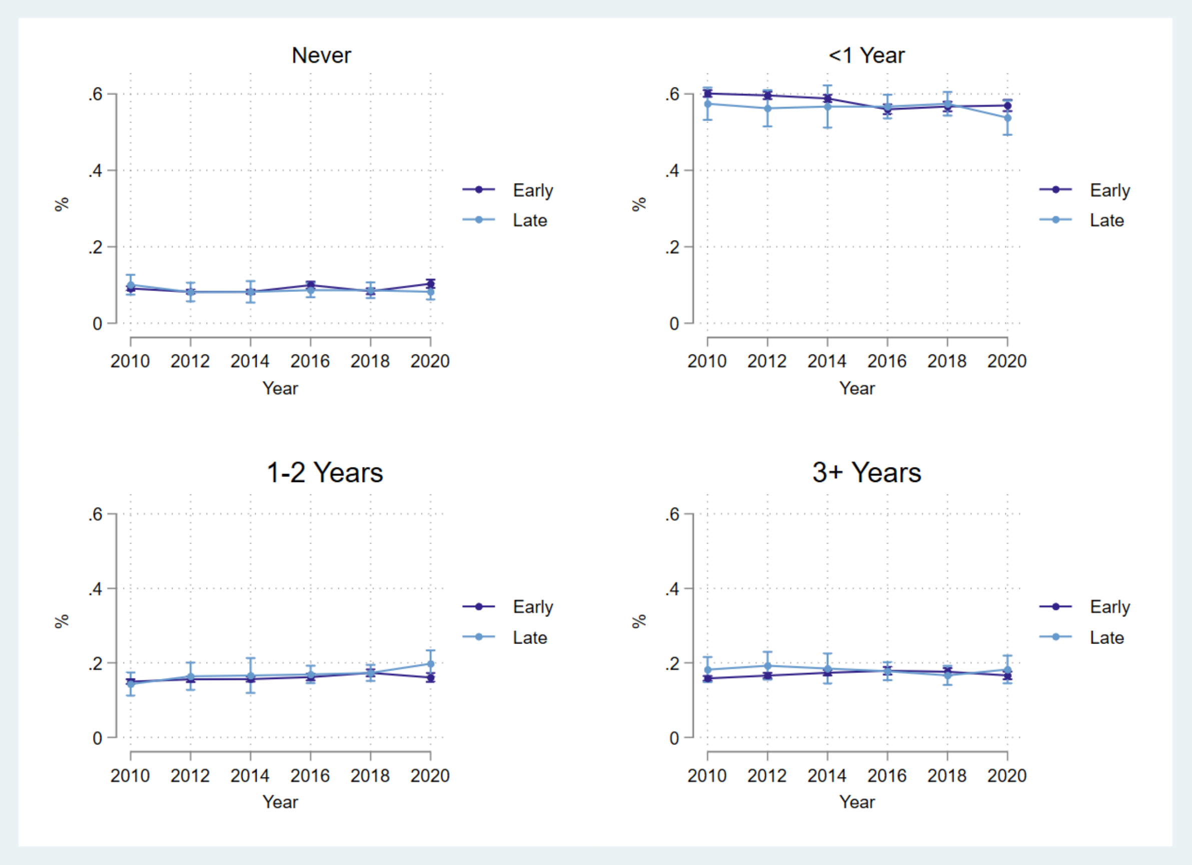


*Supplemental Figure 1 shows the year-by-year proportion of adults reporting having never received a mammogram, having received a mammogram in the past year, having received a mammogram 1-2 years ago, and having received a mammogram 3 or more years ago. The "early" cohort are adults surveyed by BRFSS in the first three months of the survey wave and the "late" cohort are adults surveyed in the final three months (Jan-Mar) or second year of the survey wave.*

##### 7.3 Supplemental Figure 3: Pap Smears

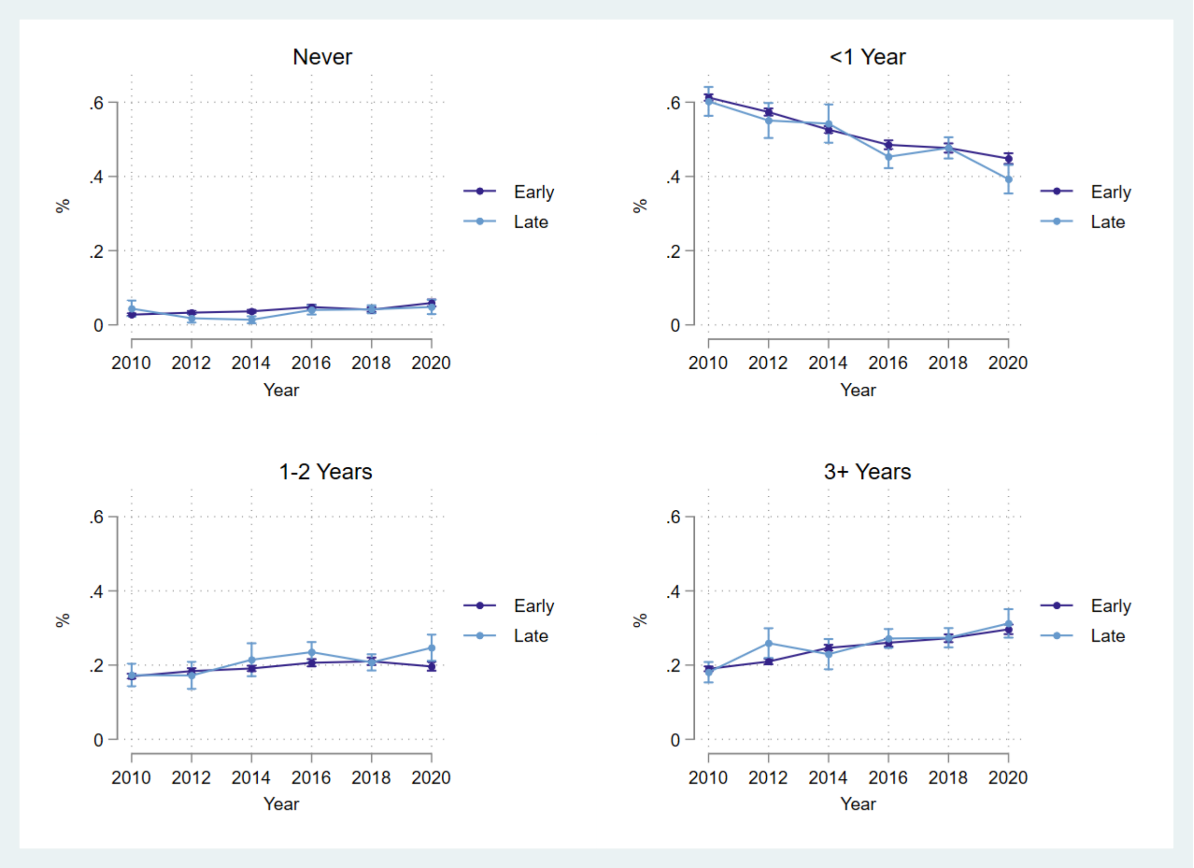


*Supplemental Figure 3 shows the year-by-year proportion of adults reporting having never received a pap smear, having received a pap smear in the past year, having received a pap smear 1-2 years ago, and having received a pap smear 3 or more years ago. The "early" cohort are adults surveyed by BRFSS in the first three months of the survey wave and the "late" cohort are adults surveyed in the final three months (Jan-Mar) or second year of the survey wave.*

#### 7.4 Supplemental Figure 4: Colonoscopies or Sigmoidoscopies

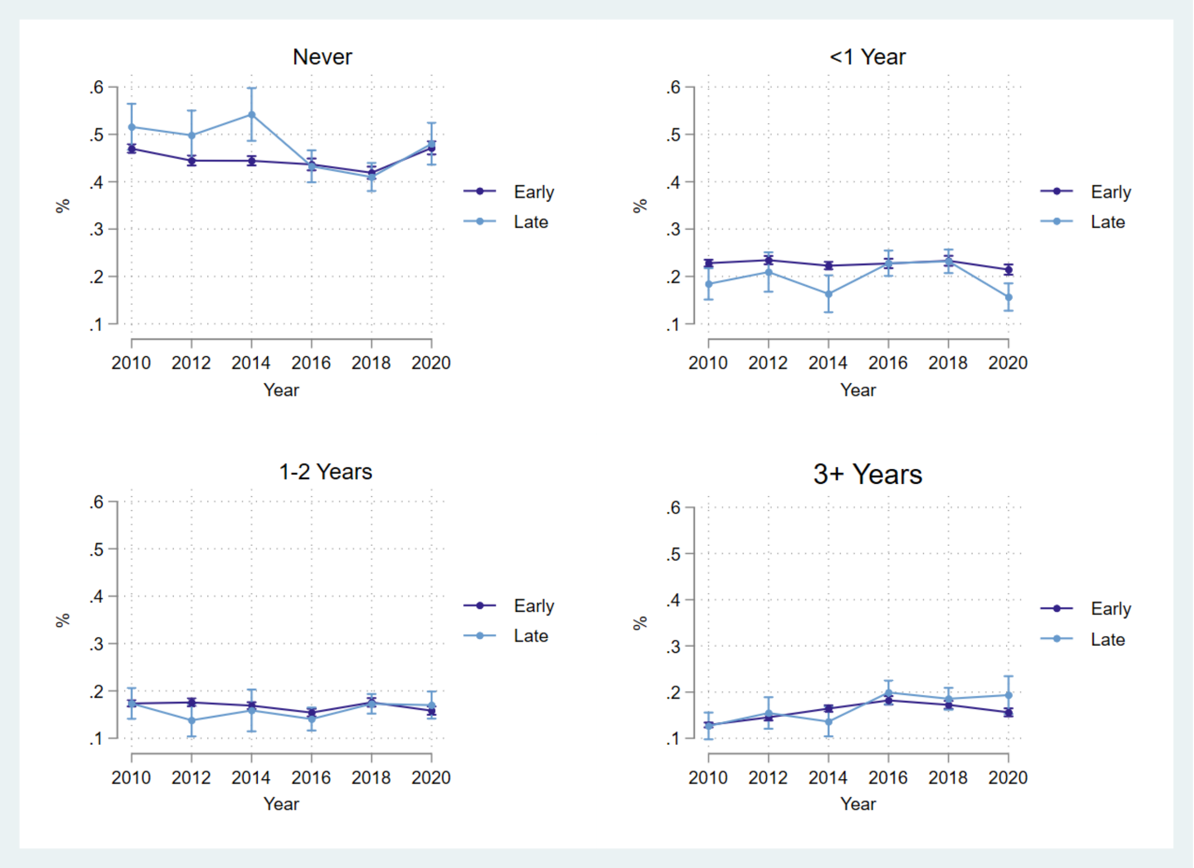


*Supplemental Figure 3 shows the year-by-year proportion of adults reporting having never received a colonoscopy or sigmoidoscopy, having received a colonoscopy or sigmoidoscopy in the past year, having received a colonoscopy or sigmoidoscopy 1-2 years ago, and having received a colonoscopy or sigmoidoscopy 3 or more years ago. The "early" cohort are adults surveyed by BRFSS in the first three months of the survey wave and the "late" cohort are adults surveyed in the final three months (Jan-Mar) or second year of the survey wave.*

Table 2: Marginal Effect Estimates for 2020*Late Group (Linear & Multinomial Logistic Regression Models)

| | | Model | |
|---|---|---|---|
| | | **Linear** | **Logistic** |
| Mammogram | Never | -0.029* (0.013) | -0.028*** (0.000) |
| | <1 Year | -0.021 (0.034) | -0.022 (0.030) |
| | 1-2 Years | 0.031* (0.015) | 0.031*** (0.000) |
| | 3+ Years | 0.020 (0.024) | 0.020 (0.037) |
| Pap Smear | Never | -0.011 (0.012) | -0.010 (0.038) |
| | <1 Year | -0.043 (0.031) | -0.055 (0.069) |
| | 1-2 Years | 0.042* (0.019) | 0.038* (0.027) |
| | 3+ Years | 0.012 (0.018) | 0.028 (0.019) |
| Colonoscopy / Sigmoidoscopy | Never | -0.010 (0.020) | -0.016 (0.019) |
| | <1 Year | -0.035* (0.016) | -0.043*** (0.001) |
| | 1-2 Years | 0.030 (0.016) | 0.038 (0.084) |
| | 3+ Years | 0.015 (0.019) | 0.021 (0.024) |

*Supplemental - Table 2 reports the 2020*Late effect estimates from our primary linear regression model (left column) and the alternative non-linear multinomail logistic regression model (right column).* $* p < 0.05$, $** p < 0.01$ $*** p < 0.001$.

Table 3: Sensitivity

| | | Mammogram | | | Pap Smear | | | Colonoscopy / Sigmoidoscopy | | |
|---|---|---|---|---|---|---|---|---|---|---|
| | | **2020*Late** | **se** | **Pre-Trend Test (p)** | **2020*Late** | **se** | **Pre-Trend Test (p)** | **2020*Late** | **se** | **Pre-Trend Test (p)** |
| *Alt. 1* | | -0.028 | (0.014) | 0.290 | -0.013 | (0.012) | 0.000^ | -0.002 | (0.020) | 0.009^ |
| *Alt. 2* | Never | -0.028 | (0.015) | 0.540 | -0.013 | (0.012) | 0.005^ | -0.002 | (0.027) | 0.081 |
| *Alt. 3* | | 0.005 | (0.006) | 0.581 | -0.013 | (0.012) | 0.108 | 0.010 | (0.011) | 0.010^ |
| *Alt. 1* | | -0.030 | (0.032) | 0.047 | -0.055 | (0.031) | 0.110 | -0.042* | (0.016) | 0.356 |
| *Alt. 2* | <1 Year | -0.030 | (0.028) | 0.368 | -0.055* | (0.025) | 0.612 | -0.042* | (0.020) | 0.294 |
| *Alt. 3* | | -0.047*** | (0.012) | 0.083 | -0.055* | (0.025) | 0.008^ | -0.043*** | (0.009) | 0.014 |
| *Alt. 1* | | 0.036* | (0.016) | 0.854 | 0.050* | (0.021) | 0.157 | 0.026 | (0.016) | 0.207 |
| *Alt. 2* | 1-2 Years | 0.036 | (0.023) | 0.935 | 0.050* | (0.022) | 0.338 | 0.026 | (0.019) | 0.331 |
| *Alt. 3* | | 0.031*** | (0.009) | 0.398 | 0.050* | (0.022) | 0.933 | 0.035*** | (0.009) | 0.416 |
| *Alt. 1* | | 0.022 | (0.022) | 0.366 | 0.017 | (0.017) | 0.011^ | 0.019 | (0.020) | 0.001^ |
| *Alt. 2* | 3+ Year | 0.022 | (0.024) | 0.539 | 0.017 | (0.025) | 0.275 | 0.019 | (0.023) | 0.260 |
| *Alt. 3* | | 0.011 | (0.009) | 0.618 | 0.017 | (0.025) | 0.056 | -0.002 | (0.009) | 0.085 |

*Supplemental - Table 3 reports the 2020*Late effect estimates for three alternative linear probability regression models. Alt 1 removes the state-level fixed effects. Alt 2 removes the state-level fixed effects and does not cluster robust standard errors at the state-level, opting only to estimate unclustered robust standard errors. Alt 3 removes fixed effects, does not cluster robust standard errors, and does not weight the analyses by BRFSS supplied sampling weights. * $p < 0.05$, ** $p < 0.01$ *** $p < 0.001$.*

Table 4: Self-reported rates (2010-2018) of most recent cancer screening for early and late survey groups

| | | **Survey Wave Group** | | | | | |
|---|---|---|---|---|---|---|---|
| | | **Eary** | | **Late** | | **T** | **p** |
| Mammogram | Never | 0.342 | (0.007) | 0.355 | (0.009) | 1.860 | 0.200 |
| | <1 Year | 0.400 | (0.007) | 0.379 | (0.008) | -3.000 | 0.100 |
| | 1-2 Years | 0.116 | (0.001) | 0.121 | (0.003) | 5.000 | 0.040 |
| | 3+ Years | 0.143 | (0.004) | 0.145 | (0.004) | 0.500 | 0.670 |
| Pap Smear | Never | 0.088 | (0.003) | 0.102 | (0.006) | 4.670 | 0.040 |
| | <1 Year | 0.467 | (0.008) | 0.420 | (0.009) | -5.880 | 0.030 |
| | 1-2 Years | 0.172 | (0.003) | 0.186 | (0.009) | 4.670 | 0.040 |
| | 3+ Years | 0.274 | (0.008) | 0.292 | (0.008) | 2.250 | 0.150 |
| Colon/Sigmoidoscopy | Never | 0.417 | (0.006) | 0.417 | (0.012) | 0.000 | 1.000 |
| | <1 Year | 0.211 | (0.005) | 0.195 | (0.008) | -3.200 | 0.090 |
| | 1-2 Years | 0.159 | (0.004) | 0.146 | (0.008) | -3.250 | 0.080 |
| | 3+ Years | 0.213 | (0.006) | 0.242 | (0.010) | 4.830 | 0.040 |

*Supplemental - Table 4 reports the rates for each mutually exclusive cancer screening category (as a proportion) of the total sample of adults surveyed by BRFSS in years 2010, 2012, 2014, 2016, and 2018. The rates are stratified by adults surveyed early in the survey wave and adults surveyed later in the survey wave. The t-statistic is the result of the two-sample proportion test.*

Table 5: Demographic rates (2010-2018) for early and late survey groups

| | Survey Wave Group | | | | | |
|---|---|---|---|---|---|---|
| **Demographic Group** | **Early** | | **Late** | | **T** | **p** |
| Age 18-24 | 0.126 | (0.002) | 0.134 | (0.003) | 4.000 | 0.060 |
| Age 25-29 | 0.081 | (0.002) | 0.091 | (0.006) | 5.000 | 0.040 |
| Age 30-34 | 0.094 | (0.002) | 0.103 | (0.003) | 4.500 | 0.050 |
| Age 35-39 | 0.081 | (0.001) | 0.088 | (0.003) | 7.000 | 0.020 |
| Age 40-44 | 0.089 | (0.001) | 0.088 | (0.002) | -1.000 | 0.420 |
| Age 45-49 | 0.080 | (0.001) | 0.077 | (0.001) | -3.000 | 0.100 |
| Age 50-54 | 0.097 | (0.001) | 0.094 | (0.002) | -3.000 | 0.100 |
| Age 55-59 | 0.081 | (0.001) | 0.081 | (0.003) | 0.000 | 1.000 |
| Age 60-64 | 0.080 | (0.001) | 0.077 | (0.002) | -3.000 | 0.100 |
| Age 65-69 | 0.061 | (0.001) | 0.059 | (0.003) | -2.000 | 0.180 |
| Age 70-74 | 0.048 | (0.001) | 0.043 | (0.004) | -5.000 | 0.040 |
| Age 75-79 | 0.038 | (0.001) | 0.030 | (0.001) | -8.000 | 0.020 |
| Age 80-84 | 0.043 | (0.002) | 0.034 | (0.002) | -4.500 | 0.050 |
| non-Hispanic White | 0.659 | (0.031) | 0.581 | (0.065) | -2.520 | 0.130 |
| non-Hispanic Black | 0.114 | (0.011) | 0.106 | (0.019) | -0.730 | 0.540 |
| non-Hispanic other race/ethnicity | 0.074 | (0.010) | 0.100 | (0.026) | 2.600 | 0.120 |
| Hispanic | 0.136 | (0.027) | 0.197 | (0.055) | 2.260 | 0.150 |
| No primary education | 0.002 | (0.001) | 0.003 | (0.001) | 1.000 | 0.420 |
| Primary education only | 0.041 | (0.005) | 0.053 | (0.011) | 2.400 | 0.140 |
| Some High School education, no degree | 0.092 | (0.002) | 0.093 | (0.004) | 0.500 | 0.670 |
| High School degree only | 0.285 | (0.008) | 0.270 | (0.015) | -1.870 | 0.200 |
| Some college education, no bachelor's degree | 0.305 | (0.005) | 0.308 | (0.007) | 0.600 | 0.610 |
| Bachelor's degree or more | 0.274 | (0.007) | 0.273 | (0.007) | -0.140 | 0.900 |
| Married | 0.524 | (0.005) | 0.515 | (0.009) | -1.800 | 0.210 |
| Divorced | 0.105 | (0.002) | 0.101 | (0.004) | -2.000 | 0.180 |
| Widowed | 0.066 | (0.002) | 0.059 | (0.004) | -3.500 | 0.070 |
| Seperated | 0.024 | (0.001) | 0.024 | (0.001) | 0.000 | 1.000 |
| Never Married | 0.237 | (0.005) | 0.251 | (0.010) | 2.800 | 0.110 |
| Unmarried Partner | 0.044 | (0.002) | 0.050 | (0.005) | 3.000 | 0.100 |

*Supplemental - Table 5 reports the rates each demographic group (used as independent variables in the primary analysis) of the total sample of adults surveyed by BRFSS in years 2010, 2012, 2014, 2016, and 2018. The rates are stratified by adults surveyed early in the survey wave and adults surveyed later in the survey wave. The t-statistic is the result of the two-sample proportion test.*

Table 6: Pre-Trend Tests

| | | Pre-Treatment Coefficient Estimates | | | | | | | | |
|---|---|---|---|---|---|---|---|---|---|---|
| | | 2010*Late | | 2012*Late | | 2014*Late | | 2016*Late | | Joint Test (p) |
| Mammogram | Never | -0.007 | (0.018) | 0.006 | (0.016) | -0.013 | (0.014) | -0.018 | (0.012) | 0.307 |
| | <1 Year | -0.012 | (0.023) | -0.022 | (0.024) | 0.002 | (0.025) | 0.021 | (0.021) | 0.027 |
| | 1-2 Years | -0.008 | (0.021) | 0.000 | (0.019) | 0.001 | (0.020) | -0.001 | (0.016) | 0.965 |
| | 3+ Years | 0.027 | (0.021) | 0.016 | (0.018) | 0.010 | (0.013) | -0.003 | (0.015) | 0.499 |
| Pap Smear | Never | 0.017 | (0.011) | -0.009 | (0.012) | -0.025** | (0.008) | -0.010 | (0.010) | 0.000^ |
| | <1 Year | 0.019 | (0.022) | 0.018 | (0.026) | 0.045* | (0.021) | -0.006 | (0.015) | 0.007^ |
| | 1-2 Years | -0.012 | (0.014) | -0.018 | (0.017) | 0.001 | (0.017) | 0.022 | (0.014) | 0.319 |
| | 3+ Years | -0.024 | (0.016) | 0.009 | (0.019) | -0.021 | (0.018) | -0.006 | (0.017) | 0.249 |
| Colonoscopy/ Sigmoidoscopy | Never | 0.019 | (0.017) | 0.015 | (0.019) | 0.019 | (0.019) | -0.024 | (0.018) | 0.257 |
| | <1 Year | -0.030 | (0.019) | -0.023 | (0.013) | -0.037 | (0.023) | -0.009 | (0.013) | 0.445 |
| | 1-2 Years | 0.012 | (0.020) | -0.021 | (0.015) | 0.006 | (0.015) | -0.009 | (0.013) | 0.096 |
| | 3+ Years | -0.011 | (0.010) | 0.014 | (0.011) | -0.018* | (0.008) | -0.016 | (0.009) | 0.002^ |

*Supplemental - Table 6 reports the year-by-year coefficients of the event-history analyses (excluding 2020) with year 2018 as the baseline. Significant coefficients indicate differences in screening rate levels between quasi-treatment cohorts. The Joint Test (p) column reports the p-value from the Wald test, testing if each of the reported coeffiecient jointly equals zero. The Bonferonni correction was used to account for multiple hypotheses. * $p < 0.05$, ** $p < 0.01$, *** $p < 0.001$. Note: $\wedge$ $p <$ multiple-test corrected significance level for Joint Test.*
